# "Serum Zinc and Selenium Levels in ICU Patients: Association with Disease Severity and Nutritional Status"

**DOI:** 10.1101/2025.05.21.25328055

**Authors:** Erfan Soltani, Aliasghar Vahidinia, Farshid Rahimi-Bashar, Davoud Ahmadimoghaddam

## Abstract

**Objective:** Patients hospitalized in the intensive care unit are usually under stress due to severe diseases and infections, which may lead to a decrease in the level of micronutrients in them. Among micronutrients, the assessment of serum selenium and zinc status in critically ill patients is of great importance, because these two minerals play a vital role in maintaining health and immune system function. Therefore, this study examines the status of serum zinc and selenium in adults hospitalized in the special care department of Baath Hospital in 2024.

**Material and Methods:** This cross-sectional study was conducted in Besat Hospital, Hamadan, during 2024 after approval by the Ethics Committee of Hamadan University of Medical Sciences. 72 patients hospitalized in the intensive care unit of Besat Hospital were selected through convenience sampling. Inclusion criteria for the study included being over 18 years of age and having been hospitalized in the ICU for one week. Written informed consent was obtained from patients or their companions for sample collection. Serum zinc and zinc levels and other laboratory factors were measured. Also, demographic and hospital information of the patient including name, age, and sex of the patient, date of hospitalization, cause of hospitalization, severity of the disease, nutritional method, duration of nutritional interruption, and outcome of the disease were recorded in the checklist. Analyses were performed using the nonparametric Mann-Whitney test, analysis of variance, and Spearman correlation test using Stata 17.0 software at a significance level of 0.05.

**Results:** According to the results, 70.8% of the patients were male and 29.2% were female, and the mean age of the patients was 54.90 years. 80.6% of the patients were on enteral nutrition (EN) and 15.3% were on oral nutrition. According to the results, 52.8% of the patients admitted to the ICU were discharged and 47.2% died. The mean serum zinc level was 15.74 micrograms/dL, of which 3.40% were zinc deficient. The mean serum selenium level was 19.97 nanograms/mL, and it was normal in 6.98% of the patients. Also, there was no statistically significant relationship between disease severity, hospital outcome, feeding interruption, feeding method, serum zinc and selenium levels in patients admitted to the ICU (p > 0.05). The mean serum zinc level based on the APACHE score showed that patients in the more severe stage of the disease had lower serum zinc levels. The mean serum zinc level in patients with APACHE score 20-30 was 71.56 micrograms/dL. The mean serum zinc level in enteral feeding patients was 74.38 and in oral feeding patients was 75.36 micrograms/dL. Also, the mean serum selenium level in enteral feeding patients was 97.25 and in oral feeding patients was 97.71 nanograms/mL.

**Conclusion:** The present study shows that zinc deficiency is significant in patients hospitalized to the intensive care unit but is not associated with disease severity, type of nutrition, interruption of nutrition, and final patient outcome. However, further studies with a larger sample size of patients are needed to draw definitive conclusions.

## 1. Introduction

Trace elements such as Zinc (Zn) and Selenium (Se) play essential roles in numerous physiological processes, particularly in the immune response, antioxidant defense, and cellular metabolism. These elements are crucial in maintaining homeostasis and have been linked to outcomes in critically ill patients, including those admitted to Intensive Care Units (ICUs) (1). The physiological stress caused by severe illness, sepsis, trauma, or other critical conditions can alter Zn and Se metabolism, leading to deficiencies that may exacerbate disease progression(2). Given the importance of these trace elements, understanding their serum levels in hospitalized ICU patients is of great clinical interest.

Critically ill patients frequently experience alterations in trace element levels, often due to factors such as inflammation, oxidative stress, and changes in nutritional intake (3). Zn plays a significant role in immune system function, enzymatic reactions, and wound healing, while Se is a vital component of selenoproteins involved in antioxidant defense mechanisms and inflammatory regulation. Deficiencies in these elements have been associated with poorer prognosis, prolonged hospitalization, and increased severity of illness (4). However, despite the recognized importance of Zn and Se in critically ill patients, limited studies have explored their relationship with disease severity, clinical outcomes, and nutritional strategies in ICU settings.

The present study was conducted in the ICU unit of Be’sat Hospital in Hamadan, Iran, from November to December 2024, to assess the serum levels of Zn and Se in ICU patients and explore their associations with disease severity scores (APACHE II & SOFA) and nutritional modes. The Acute Physiology and Chronic Health Evaluation (APACHE II) score (5) and the Sequential Organ Failure Assessment (SOFA) score are widely used in ICU settings to estimate disease severity and predict patient outcomes (6). Given their prognostic value, investigating whether Zn and Se levels correlate with these scores could provide insights into their role in critical illness.

This study aimed to:

1. Evaluate serum Zn and Se levels in ICU patients and compare them across different nutritional strategies (oral, enteral, and parenteral nutrition).
2. Assess the correlation between Zn/Se levels and disease severity using APACHE II and SOFA scores.
3. Determine whether Zn and Se levels predict disease severity through statistical modeling.

By addressing these objectives, this research aims to contribute valuable data to the existing literature on trace element metabolism in critically ill patients and its potential implications for clinical management.

## 2. Methods

### 2.1. Study Design and Population

This study was designed as a cross-sectional observational study conducted in the intensive care unit (ICU) of a tertiary care hospital. The study aimed to assess the relationship between serum zinc (Zn) and selenium (Se) levels and clinical severity indicators, including the Acute Physiology and Chronic Health Evaluation II (APACHE II) score and the Sequential Organ Failure Assessment (SOFA) score.

#### Inclusion Criteria

Patients admitted to the ICU were included in the study if they met the following criteria:

- Age ≥ 18 years
- Hospitalized in the ICU for at least 7 days
- Availability of serum Zn and Se measurements during ICU admission

#### Exclusion Criteria

Patients were excluded if they:

- Were under 18 years old
- Had an ICU stay of less than 7 days
- Had received zinc or selenium supplementation during hospitalization
- Had pre-existing conditions known to influence trace element levels, such as chronic liver disease, chronic kidney disease, or known malabsorption disorders

### 2.2. Data Collection

After obtaining informed consent from patients or their legal guardians, clinical and demographic data were collected from electronic medical records. The following variables were recorded:

#### Demographic and Clinical Data

- Age (years)
- Gender (Male/Female)
- ICU Admission Diagnosis (e.g., sepsis, trauma, post-surgical recovery)
- Length of ICU Stay (days)

#### Nutritional Mode

Patients were classified based on their primary mode of nutrition during ICU admission into three groups:

1. Oral Nutrition (ON): Patients who were able to consume a normal diet or modified oral diet.
2. Enteral Nutrition (EN): Patients receiving tube feeding through nasogastric or percutaneous endoscopic gastrostomy (PEG) tubes.
3. Total Parenteral Nutrition (TPN): Patients receiving intravenous nutrition, bypassing the digestive tract.

#### Clinical Severity Scores

- APACHE II Score: A standardized scoring system assessing disease severity, calculated based on physiological variables recorded within 24 hours of ICU admission.
- SOFA Score: An organ dysfunction assessment tool based on respiratory, cardiovascular, hepatic, coagulation, renal, and neurological parameters.

### 2.3. Blood Sample Collection and Analysis

Serum Zn and Se levels were measured from blood samples obtained within 7 days of ICU admission or more.

#### Zinc Measurement

- Blood samples were collected in trace-element-free vacutainers to prevent contamination.
- Serum Zn concentrations were analyzed using 5-Br-PAPS colorimetric assay, a validated method for detecting Zn levels in clinical settings (7).
- Results were reported in micrograms per deciliter (μg/dL).

#### Selenium Measurement

- Samples for Se analysis were centrifuged and stored at -80°C before processing.
- Se concentrations were determined using Inductively Coupled Plasma Mass Spectrometry (ICP-MS), which offers high precision for trace element detection (8)
- Results were reported in micrograms per liter (ng/mL).

### 2.4. Statistical Analysis

All statistical analyses were performed using Stata 17.0 software.

#### Descriptive Statistics

- Continuous variables (Zn, Se, APACHE II, SOFA scores) were summarized using mean ± standard deviation (SD).
- Categorical variables (gender, nutrition type) were summarized using percentages and frequencies.

Comparison of Zn and Se Levels Across Nutrition Groups

#### Correlation Analysis

To evaluate the relationship between Zn/Se levels and disease severity, Pearson correlation coefficients (r-values) were computed:

- Zn vs. APACHE II score
- Zn vs. feeding interruption duration
- Zn vs. patient’s clinical outcome
- Zn vs. SOFA score
- Zn vs. NUTRIC score
- Se vs. APACHE II score
- Se vs. feeding interruption duration
- Se vs. patient’s clinical outcome
- Se vs. SOFA score
- Se vs. NUTRIC score
- Zn vs. Se

Statistical significance was set at p < 0.05.

#### Multiple Regression Analysis

To assess whether Zn and Se levels could predict disease severity, we performed multiple linear regression with APACHE II and SOFA scores as dependent variables. Independent variables included:

- Serum Zn levels
- Serum Se levels
- Age (as a confounding factor)

#### 2.5. Ethical Considerations

- The study was approved by the Institutional Review Board (IRB).
- Informed consent was obtained from all participants or their legal representatives.
- Data was anonymized to protect patient confidentiality. Conclusion of Methods Section

Our methodology incorporated a robust statistical approach, ensuring rigorous evaluation of potential associations between Zn/Se levels and ICU disease severity. The inclusion of Stata 17.0, correlation, multiple regression, and polynomial regression analyses allowed us to comprehensively explore both linear and nonlinear relationships in our dataset.

## 3. Results

This section presents the findings of our study, including patient demographics, trace element levels (Zn and Se), statistical analyses exploring differences across nutritional groups, and correlations with disease severity scores. We employed a comprehensive statistical approach, including ANOVA, t-tests, correlation analysis, multiple regression, and polynomial regression, to investigate the relationships between Zn/Se levels and clinical parameters.

### 3.1. Patient Characteristics

A total of 72 ICU patients were included in this study. The demographic and clinical characteristics are summarized in table 1.

- Mean Age: 54.9 ± 20.24 years
- Gender Distribution: 51 males (70.8%) and 21 females (29.2%)
- Nutritional Mode Distribution:

`o` Oral Nutrition (ON): 11 patients (15.3%)
`o` Enteral Nutrition (EN): 58 patients (80.6%)
`o` Total Parenteral Nutrition (TPN): patients (0%)

Notice: 3 patients were NPO(4.2%) regiment when we conducted our samples

- Mean APACHE II Score: 18.5 ± 7.2
- Mean SOFA Score: 7.9 ± 3.6

**Table. 1.**

| Percent | Number of patients | Cause of admission | Percent | Number of patients | Cause of admission |
| --- | --- | --- | --- | --- | --- |
| 2.8 | 2 | Lung cancer | 1.4 | 1 | abdominal hernia |
| 16.7 | 12 | MPT | 1.4 | 1 | Aortic aneurysm |
| 1.4 | 1 | neck trauma | 1.4 | 1 | cancer |
| 2.8 | 2 | Perforating peptic ulcer | 2.8 | 2 | Cerebral edema |
| 1.4 | 1 | peritonitis | 1.4 | 1 | cerebral hematoma |
| 1.4 | 1 | pleural efusion | 2.8 | 2 | Cerebral tumor |
| 2.8 | 2 | Pneumonia | 1.4 | 1 | Cervical fracture |
| 1.4 | 1 | POD | 1.4 | 1 | Cholelithiasis |
| 1.4 | 1 | post surgery infection | 1.4 | 1 | Coleostomy |
| 2.8 | 2 | hydrocephalia | 1.4 | 1 | Colon cancer |
| 6.9 | 5 | ICH | 1.4 | 1 | COPD |
| 2.8 | 2 | intestinal obstruction | 1.4 | 1 | Craniotomia |
| 1.4 | 1 | left hemothoraxia | 4.2 | 3 | CVA |
| 1.4 | 1 | loss of conscience | 1.4 | 1 | DKA |
| 1.4 | 1 | PTE | 1.4 | 1 | esophagus rupture |
| 1.4 | 1 | right hemiplastic aphasia | 1.4 | 1 | F.D |
| 1.4 | 1 | S.M | 1.4 | 1 | F.x C5&C6 |
| 6.9 | 5 | SAH | 1.4 | 1 | f.x T2&pelvic |
| 2.8 | 2 | SDH | 1.4 | 1 | frontal hematoma |
| 1.4 | 1 | Toxicity | 1.4 | 1 | Gastric bleeding |
| 1.4 | 1 | Weakness | 1.4 | 1 | gastric cancer |
|  |  |  | 1.4 | 1 | Hernia |

**Figure 1.**
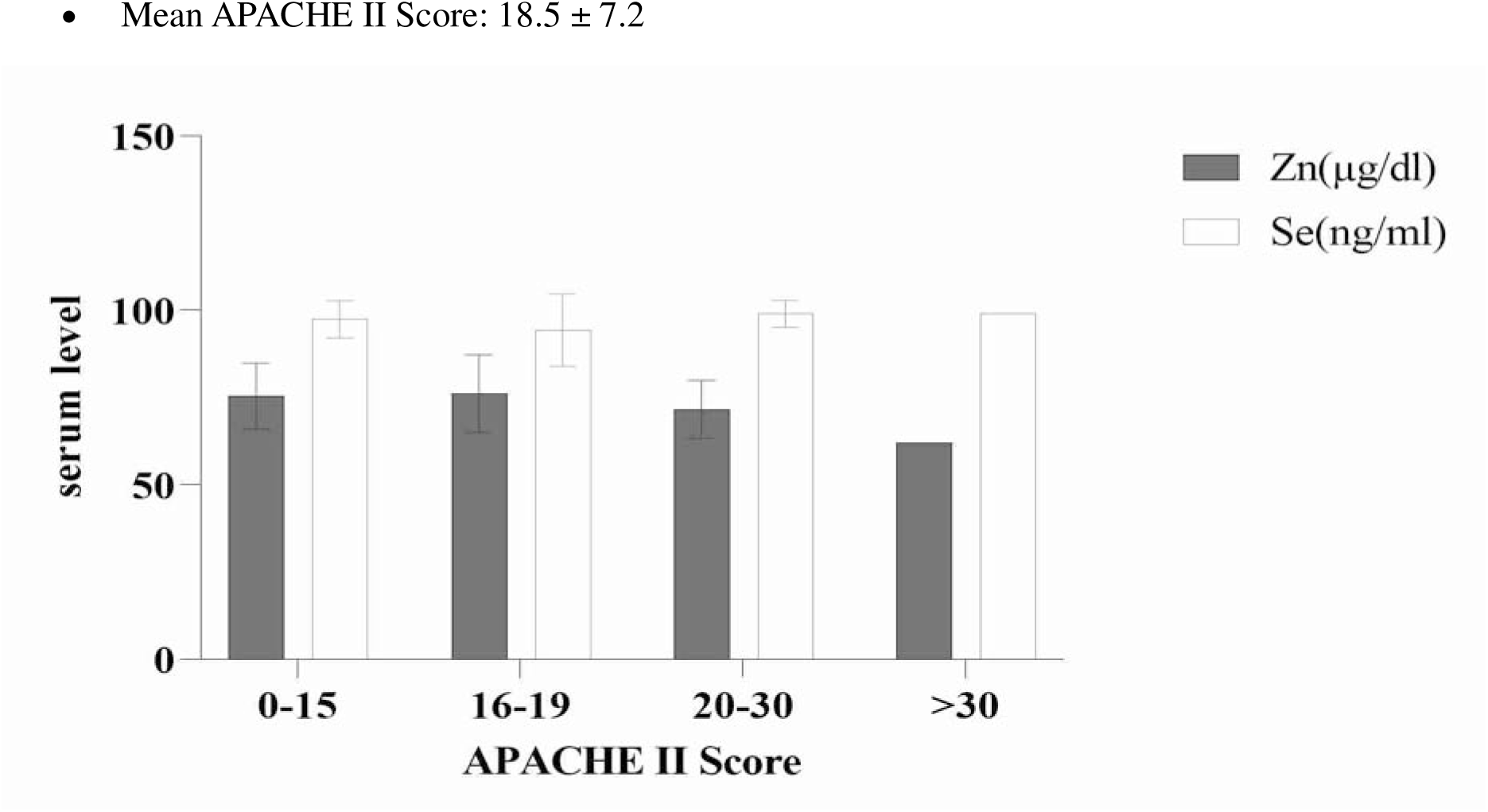
Distribution of ICU Patients by APACHE II Score.

**Figure 2.**
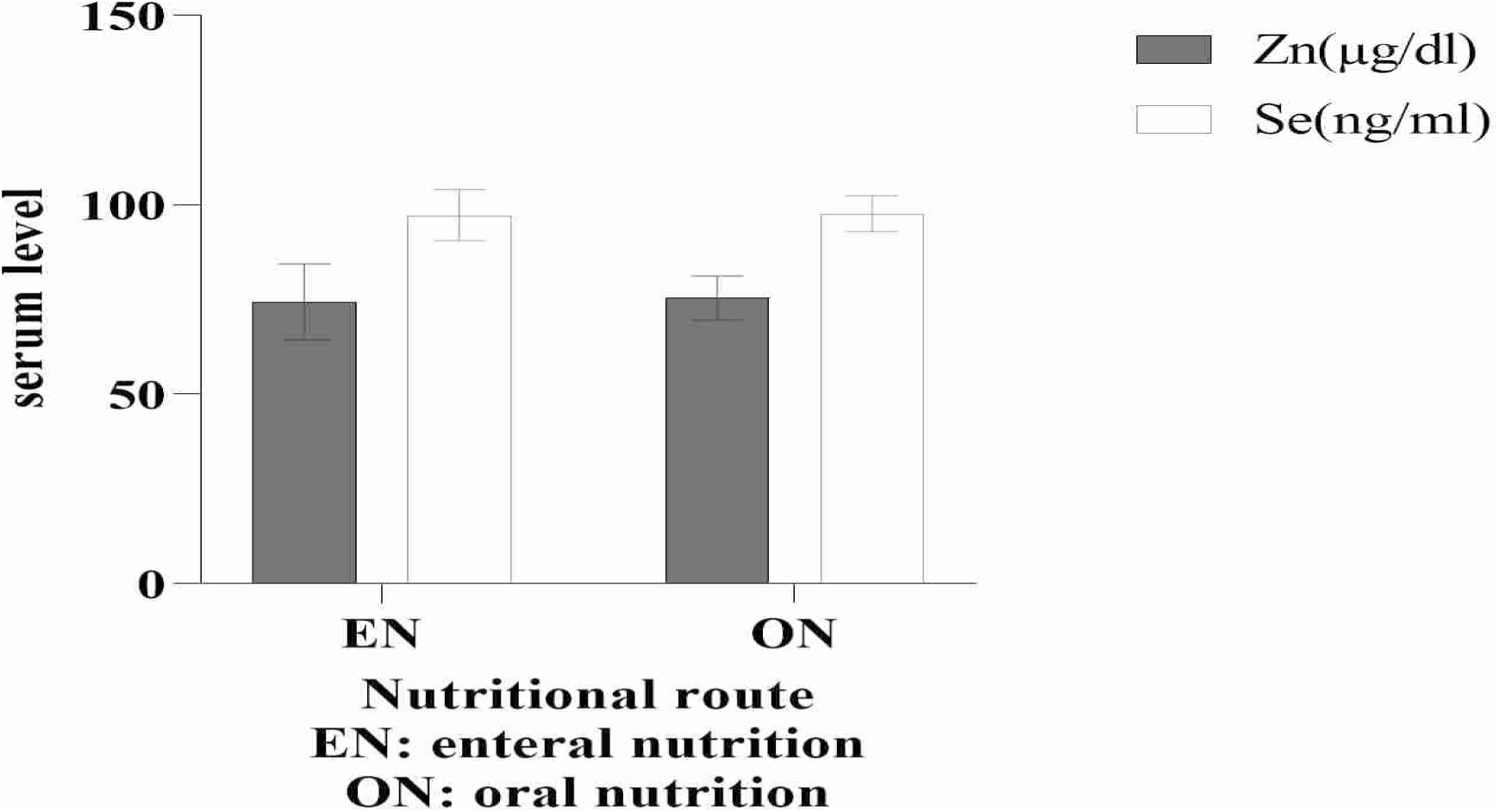
Distribution of ICU patients by Nutrition route.

**Figure 3.**
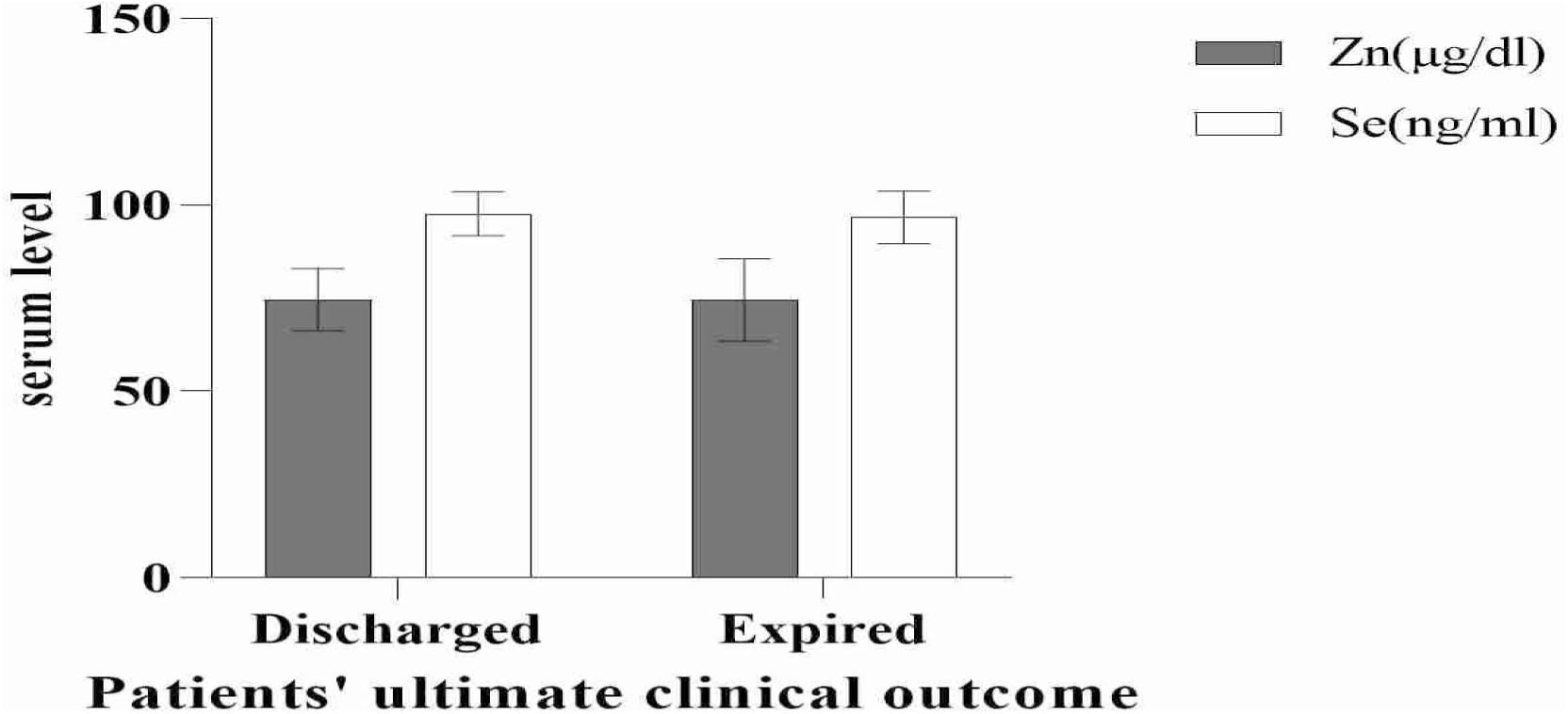
Patients’ Ultimate Clinical Outcome.

**Figure 4.**
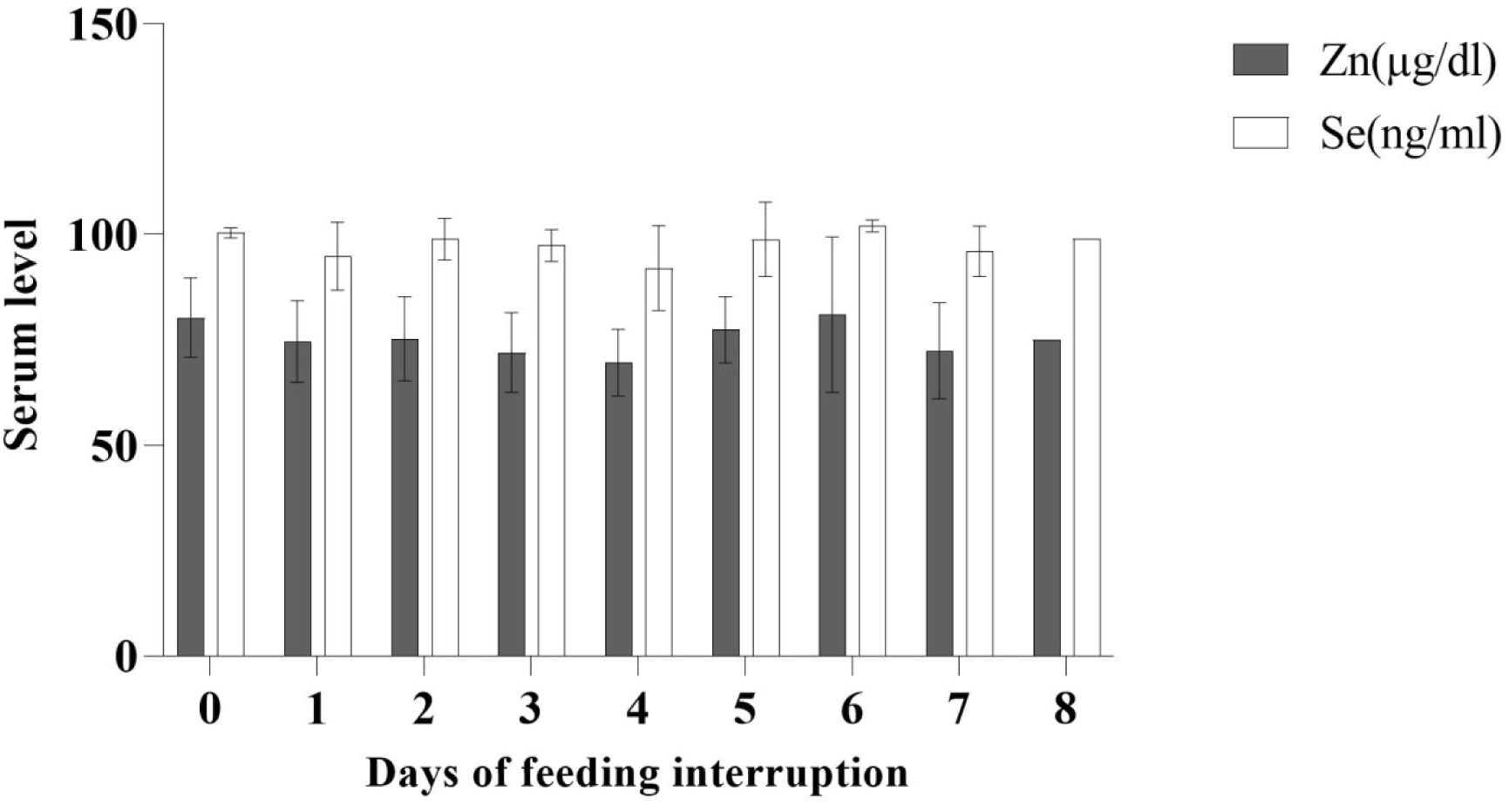
Distribution ICU Patients by Feeding Interruption Duration (days)

### 3.2. Zinc and Selenium Levels

To assess whether Zn and Se levels varied based on the type of nutritional intake (Oral, EN, or TPN), we conducted a one-way ANOVA test.

- Zinc Levels (Mean ± SD, μg/dL): 74.15 ± 9.64 (minimum:51-maximum:94.1)

According to the Normal range for serum Zn level 40.3% of patients had insufficiency and rest were within normal range.

- Selenium Levels (Mean ± SD, ng/mL): 97.19 ± 6.44 (minimum:70-maximum:113.3)
- According to the Normal range for serum Se level 1.4% of patients had insuffiency and the rest were within normal range

Interpretation: The results indicate that there were no significant differences in Zn and Se serum levels across different nutrition types. This suggests that nutritional intake mode may not be the primary determinant of Zn and Se concentrations in ICU patients.

### 3.3. Correlation Analysis: Zn, Se, and Disease Severity

To explore potential relationships between Zn/Se levels and clinical severity indices, we performed Pearson correlation analysis between Zn and Se levels and APACHE II and SOFA scores. The results are shown in Table 3.

- Zinc vs. APACHE II: p-value = 0.081 (No significant correlation)
- Zinc vs. SOFA: r = −0.07, p = 0.36 (No significant correlation)
- Selenium vs. APACHE II: r = −0.08, p = 0.30 (No significant correlation)
- Selenium vs. SOFA: r = −0.05, p = 0.47 (No significant correlation)
- Zinc vs. Selenium: r = 0.12, p = 0.16 (Weak positive correlation, not significant)

Interpretation: No significant correlations were found between Zn/Se levels and disease severity scores. This indicates that Zn and Se serum levels do not strongly predict APACHE II or SOFA scores in this ICU population.

### 3.4. Regression Analysis: Predicting Disease Severity from Zn and Se Levels

To determine whether Zn and Se levels are predictors of disease severity, we performed multiple linear regression analysis using APACHE II and SOFA scores as dependent variables.

Interpretation: The regression models suggest that Zn and Se levels are not significant predictors of disease severity as measured by APACHE II or SOFA scores. Even when accounting for patient age, the explanatory power of these models remains very low.

### 3.5. Nonlinear Analysis (Polynomial Regression Models)

Given that biological relationships may not always be linear, we conducted polynomial regression analysis to explore potential non-linear associations between Zn/Se levels and disease severity scores.

### 3.6. Summary of Key Findings

1. Zn and Se levels do not significantly vary across different nutrition types (oral, enteral, parenteral).
2. No significant correlations were found between Zn/Se levels and APACHE II or SOFA scores.
3. Zn and Se are not significant predictors of disease severity in multiple linear regression models.
4. Polynomial regression confirms the weak explanatory power of Zn and Se in predicting disease severity scores.

### 3.7. Visualizations

To illustrate our findings, we generated the following plots:

1. Boxplots of Zn and Se levels by nutrition type, confirming no significant differences.
2. Scatter plots of Zn/Se vs. APACHE II and SOFA scores, showing no clear trend.
3. Regression plots, visualizing weak/non-existent associations. Conclusion of Results Section

Overall, our findings indicate that Zn and Se levels do not significantly influence disease severity in ICU patients. These results suggest that other factors, such as inflammation, metabolic stress, and organ dysfunction, may play a more significant role in determining disease severity than serum Zn and Se levels alone. Further research should explore additional biomarkers and longitudinal trends in Zn and Se levels over the course of ICU admission.

**Figure 5.**
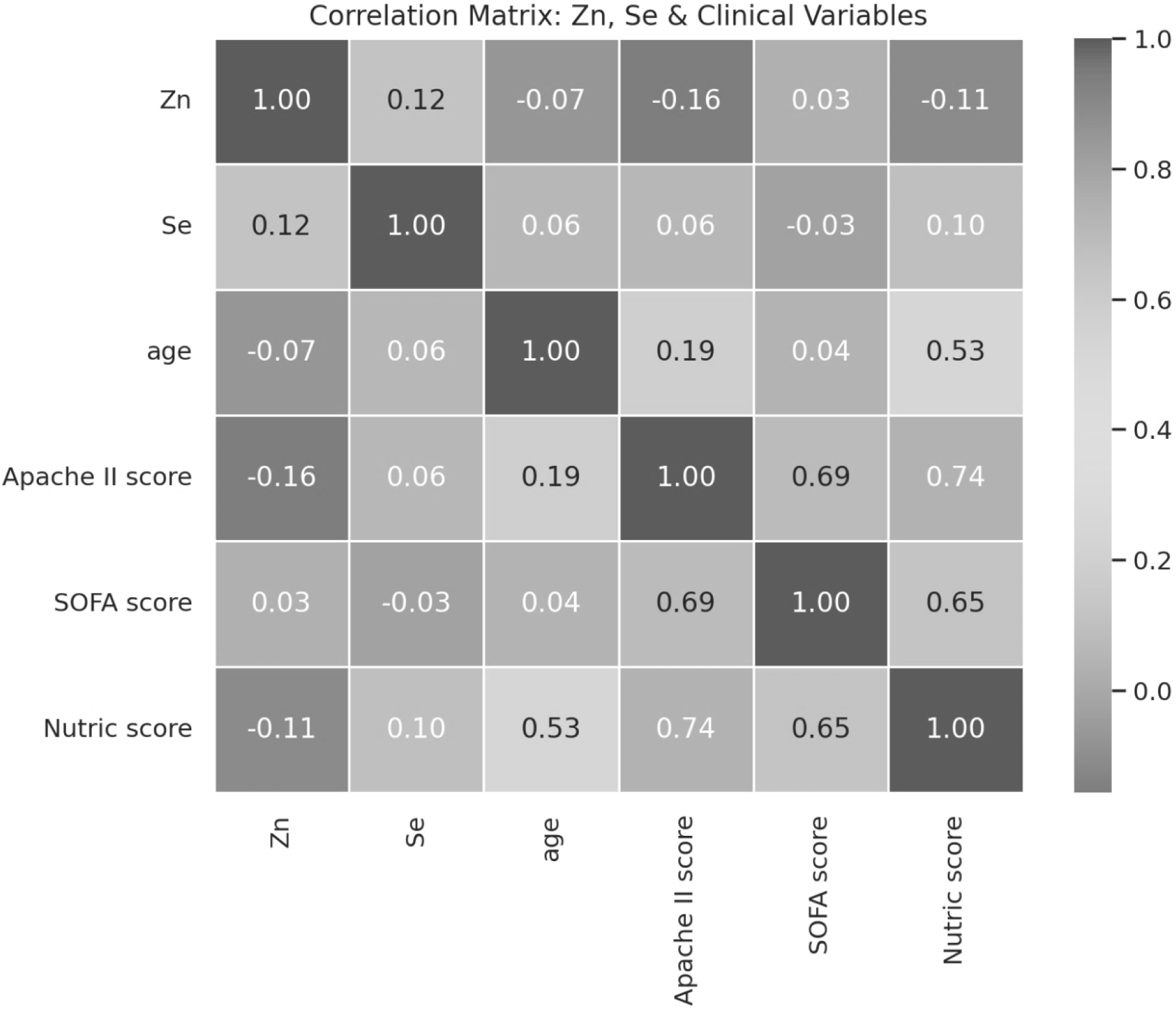
Correlation matrix: Zinc, Selenium and clinical variables.

**Figure 6.**
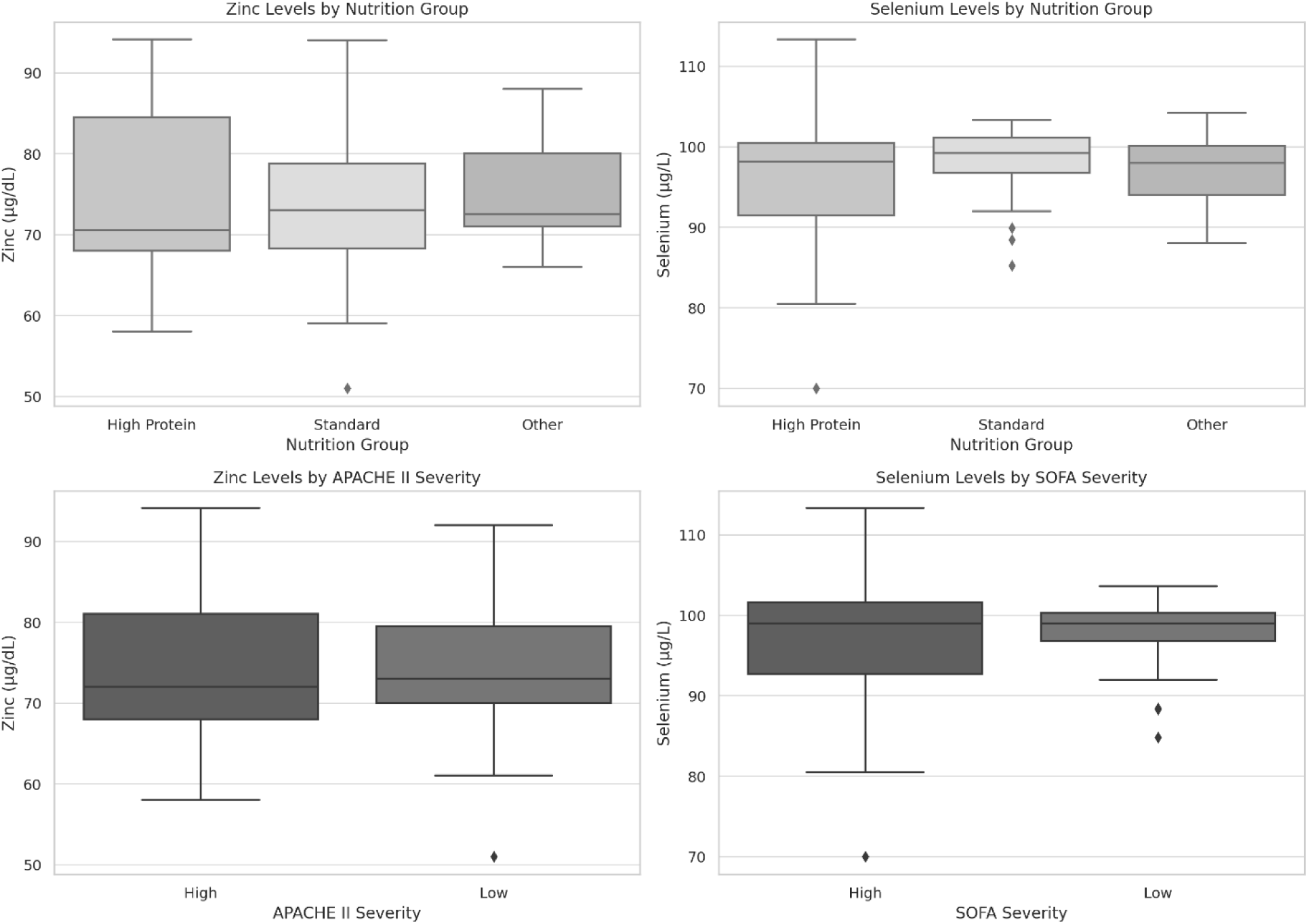
Correlation between trace elements and clinical variables.

## 4. Discussion

The present study sought to investigate the association between zinc (Zn) and selenium (Se) serum levels and clinical severity among critically ill patients admitted to the ICU. Our findings indicate that Zn and Se levels do not significantly vary based on the type of nutrition (oral, enteral, or parenteral) and do not show a meaningful correlation with disease severity indices such as APACHE II and SOFA scores. These results suggest that the physiological mechanisms regulating Zn and Se levels in critically ill patients may not be solely dependent on nutritional intake but rather influenced by complex metabolic and inflammatory processes.

### 4.1. The Role of Zinc and Selenium in Critical Illness

Zn and Se are essential trace elements with significant roles in immune function, antioxidant defense, and inflammatory regulation (1). Zn is a crucial cofactor for over 300 enzymes involved in cellular proliferation, protein synthesis, and wound healing, whereas Se is integral to the function of selenoproteins, which exhibit antioxidant properties and regulate immune responses (9). Deficiencies in these elements have been associated with impaired immune function, increased oxidative stress, and poor clinical outcomes in critically ill patients (4).

Previous studies have reported that Zn and Se deficiencies are common in ICU patients, particularly those with sepsis, systemic inflammation, or organ failure (10). However, in our study, Zn and Se levels were not significantly associated with the severity of illness as measured by APACHE II or SOFA scores. This discrepancy may be due to differences in patient populations, measurement techniques, or the timing of trace element assessments relative to disease progression.

### 4.2. Zn and Se Levels Across Different Nutrition Groups

One of the key objectives of this study was to determine whether the method of nutritional intake—oral, enteral (EN), or total parenteral nutrition (TPN)—influences Zn and Se serum levels. Contrary to our initial hypothesis, there were no significant differences in Zn and Se concentrations among these groups. This finding is consistent with prior research indicating that Zn and Se homeostasis in critically ill patients is regulated by factors beyond mere dietary intake (11).

During critical illness, trace element levels can be influenced by shifts in metabolic demand, redistribution between compartments, and interactions with inflammatory mediators (12). Inflammatory responses lead to increased Zn sequestration in the liver and reduced plasma Zn levels due to upregulation of metallothioneins (13). Similarly, Se concentrations may decline during acute illness due to increased utilization by selenoproteins and alterations in protein-binding dynamics (14). These mechanisms may explain why Zn and Se levels in our study were not significantly altered by the type of nutrition received.

### 4.3. Correlation Between Zn, Se, and Disease Severity

In contrast to studies that have linked Zn and Se deficiencies to worse clinical outcomes (4), we found no strong correlation between Zn/Se levels and APACHE II or SOFA scores. Previous investigations have suggested that low Zn and Se levels might be indicative of prolonged ICU stays, organ dysfunction, and increased mortality(15). However, our results suggest that these trace elements alone may not be reliable biomarkers for predicting disease severity in all ICU populations.

One potential explanation is that Zn and Se depletion occurs in a time-dependent manner. Early during ICU admission, Zn and Se levels may not reflect the full extent of metabolic stress, whereas prolonged ICU stays may lead to progressive depletion(16). Additionally, genetic variability in trace element metabolism and individual differences in baseline nutritional status could contribute to the variability observed in Zn and Se levels (1).

### 4.4. Clinical Implications and Potential Future Directions

Our findings suggest that while Zn and Se play important physiological roles in critical illness, their serum concentrations may not be reliable indicators of disease severity or nutritional adequacy. This has important implications for clinical practice, as routine Zn and Se monitoring may not be necessary for all ICU patients unless there is a clear indication of deficiency.

Future research should focus on:

1. Longitudinal Studies: Examining Zn and Se levels over time to determine whether progressive depletion is associated with worsening clinical outcomes.
2. Interventional Trials: Assessing whether Zn and Se supplementation improves outcomes in specific ICU subpopulations, such as those with sepsis or multi-organ failure (17).
3. Biomarker Integration: Investigating the role of Zn and Se in conjunction with other inflammatory and metabolic markers to create a more comprehensive picture of nutritional status in critically ill patients.

### 4.5. Strengths and Limitations

A major strength of this study is its comprehensive assessment of Zn and Se levels in a well-defined ICU cohort, along with detailed statistical analyses to explore their associations with nutrition type and disease severity. However, several limitations should be acknowledged:

- Single-Time Measurement: Zn and Se levels were assessed at a single time point, limiting our ability to track changes over time.
- Potential Confounders: The presence of comorbid conditions, inflammatory status, and medication use may have affected Zn and Se metabolism, introducing potential confounding effects.

## 5. Conclusion

This study found no significant association between Zn/Se serum levels and ICU disease severity scores, nor any significant differences based on nutrition type. These findings highlight the complexity of trace element regulation in critically ill patients and suggest that Zn and Se alone may not serve as standalone biomarkers for disease severity. Further research is needed to explore their role in ICU patient management, particularly in the context of dynamic metabolic changes and individualized supplementation strategies.

## Data Availability

All data produced in the present work are contained in the manuscript

